# Efficacy and Safety of Remdesivir for COVID-19 Treatment: An Analysis of Randomized, Double-Blind, Placebo-Controlled Trials

**DOI:** 10.1101/2020.06.22.20136531

**Authors:** Yun Zhu, Zhaowei Teng, Lirong Yang, Shuanglan Xu, Jie Liu, Yirong Teng, Qinggang Hao, Dake Zhao, Xiaolan Li, Sheng Lu, Yong Zeng

**Author notes:** All authors contributed equally to this paper. **Correspondence to:** Yun Zhu, The Sixth Affiliated Hospital of Kunming Medical University, 21 Nieer Road, Yuxi 653100, Yunnan, China.

## Abstract

**BACKGROUND:** Remdesivir, an inhibitor of viral RNA-dependent RNA polymerases, has been identified as a candidate for COVID-19 treatment. However, the therapeutic effect of remdesivir is controversial.

**METHODS:** We searched PubMed, Embase, and the Cochrane Central Register of Controlled Trials, from inception to June 11, 2020 for randomized controlled trials on the clinical efficacy of remdesivir. The main outcomes were discharge rate, mortality, and adverse events. This study is registered at INPLASY (INPLASY202060046).

**RESULTS:** Data of 1075 subjects showed that remdesivir significantly increased the discharge rate of patients with COVID-19 compared with the placebo (50.4% vs. 45.29%; relative risk [RR] 1.19 [95% confidence interval [CI], 1.05–1.34], I^2^ = 0.0%, P = 0.754). It also significantly decreased mortality (8.18% vs. 12.70%; RR 0.64 [95% CI, 0.44–0.92], I^2^ = 45.7%, P = 0.175) compared to the placebo. Data of 1296 subjects showed that remdesivir significantly decreased the occurrence of serious adverse events (RR 0.77 [95% CI, 0.63–0.94], I^2^ = 0.0%, P = 0.716).

**CONCLUSION:** Remdesivir is efficacious and safe for the treatment of COVID-19.

**TRIAL REGISTRATION NUMBER:** This study is registered at the International Platform of Registered Systematic Review and Meta-analysis Protocols (INPLASY202060046).

---

In December 2019, in Wuhan, China, several cases of pneumonia were caused by a novel coronavirus, the severe acute respiratory syndrome coronavirus 2 (SARS-CoV-2). The World Health Organization (WHO) officially designated this new disease as coronavirus disease 2019 (COVID-19).^1,2^ COVID-19 is declared a global pandemic, and has led to over 8.5 million infections and 452 thousand deaths up to now. Due to the lack of a specific antiviral agent that targets the virus, COVID-19 pandemic remains largely uncontrolled with high morbidity and mortality, posing a huge challenge globally.^3^ Therefore, it is of great importance to explore safe and effective treatment options.

Remdesivir (GS-5734) is an inhibitor of the viral RNA-dependent RNA polymerase and may cause premature RNA-chain termination. It has a broad antiviral spectrum that covers pneumoviruses, paramyxoviruses, filoviruses, and coronaviruses. Furthermore, remdesivir plays an important role in the treatment of Middle East respiratory syndrome (MERS-CoV).^4^ Interestingly, remdesivir is effective in inhibiting SARS-CoV-2 growth *in vitro*;^5^ it has reportedly reduced viral load in the lungs and improved disease symptoms in a mouse model of SARS-CoV.^6^ It has been identified by the WHO as the most promising therapeutic candidate for the treatment of COVID-19.^7^ However, since the therapeutic effect of remdesivir is controversial,^7-10^ we aimed to evaluate clinically meaningful evidence on its efficacy and safety in the treatment of COVID-19 via an analysis of pooled randomized controlled trials (RCTs).

## Methods

### SEARCH STRATEGY AND CRITERIA

We systematically searched PubMed, Embase, and the Cochrane Central Register of Controlled Trials from their inception to June 11, 2020, to identify RCTs on remdesivir treatment for COVID-19, with no restrictions on publication type or language. Our main search keywords were “coronavirus disease 2019,” “COVID-19,” “novel coronavirus 2019,” “2019 novel coronavirus,” “severe acute respiratory syndrome-corona virus-2,” “SARS-CoV-2,” “remdesivir,” “gs 5734,” and “randomized controlled trial” (Appendix 1). Studies were considered eligible if they met the following criteria: (1) presented original data from a randomized, double-blind, placebo-controlled trial; (2) used two comparator groups with one receiving remdesivir and the other receiving a placebo; (3) reported the number of patients discharged from hospital, death, and adverse events as outcomes; and (4) had adequate data to be pooled for the analysis. Two authors (TZW and ZY) independently selected eligible articles. First, titles and abstracts were evaluated to exclude non-relevant articles. Then, full text was read to confirm whether studies conformed to our inclusion criteria. Disputes between the two authors were resolved by consensus. The quality of the studies was assessed using the Jadad scoring system,^11^ in which the maximum score is 5. We defined high quality as a Jadad score of ≥3.0.

### DATA ANALYSIS

Two authors (ZY and TZW) extracted all data. For each study, we recorded the following variables: first author’s last name, publication year, sample size, participants’ sex and age, and patient distribution on a six-category scale (Appendix 2) at enrollment and at 15 ± 2 days after enrollment, adverse events, and outcome measurements related to risk estimates with 95% confidence interval (CI).

Pre-specified endpoints included discharge and death as primary endpoints with adverse events including anemia, pulmonary embolism, deep vein thrombosis, acute kidney injury, septic shock, and cardiac arrest as key secondary endpoints. Each outcome was assessed according to the definitions reported in the study protocols. We conducted a pooled analysis to measure the efficacy and safety of remdesivir for COVID-19 treatment. We pooled the risk ratios (RR) and 95% CI values calculated using chi-squared (χ^2^) tests. Cochran Q and I^2^ statistics were used to evaluate statistical heterogeneity.^12^ When the p-value was >0.1 and the I^2^ value was <50%, a fixed-effects model^13^ was used to estimate the overall summary effect sizes. When either the p-value was <0.1 or the I^2^ value was >50%, the data were considered to be heterogeneous, and a random-effects model^14^ was applied to determine the overall summary effect sizes. STATA software v12.0 (College Station, TX, USA) was used to analyze the data.

### TRIAL REGISTRATION

This study is registered at the International Platform of Registered Systematic Review and Meta-analysis Protocols (INPLASY202060046).

## Results

We found 87 relevant studies through our initial search. After removing 10 duplicate articles and 21 reviews, 56 remained. Eventually, two studies^7-8^ were finally included in our analysis after screening titles and abstracts. No eligible studies were identified by screening the bibliographies of the relevant studies (Figure 1). According to the Jadad scale, two studies were of high quality with scores of 5 (Table 1). A total of 1299 patients with COVID-19 randomly allocated to a group treated with remdesivir (n = 699) or group treated with placebo (n = 600) were included in our analysis. The distribution of patients based on a six-category scale at enrollment are shown in Table 1, Figure 2A, and Figure 2B. Among the included patients, the average age was over 50 years and number of men exceeded that of women. Details of the characteristics of the studies are summarized in Table 1.

**Table 1.**
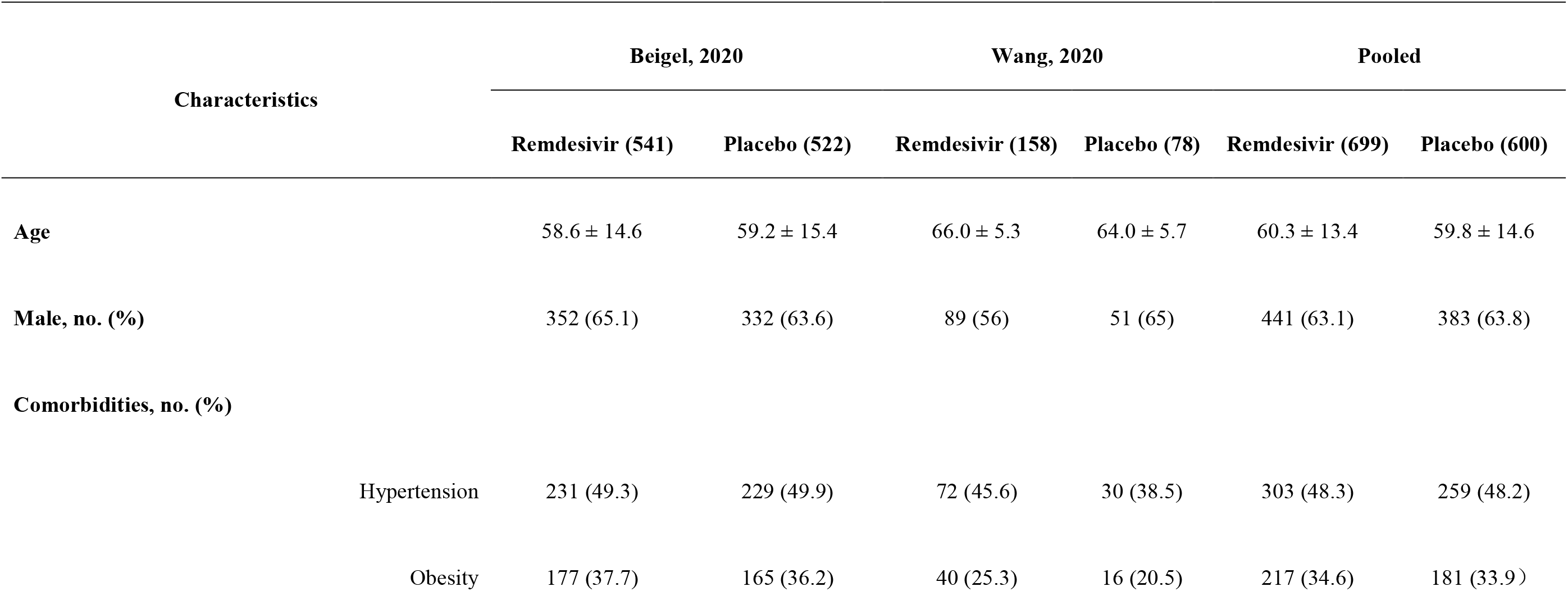
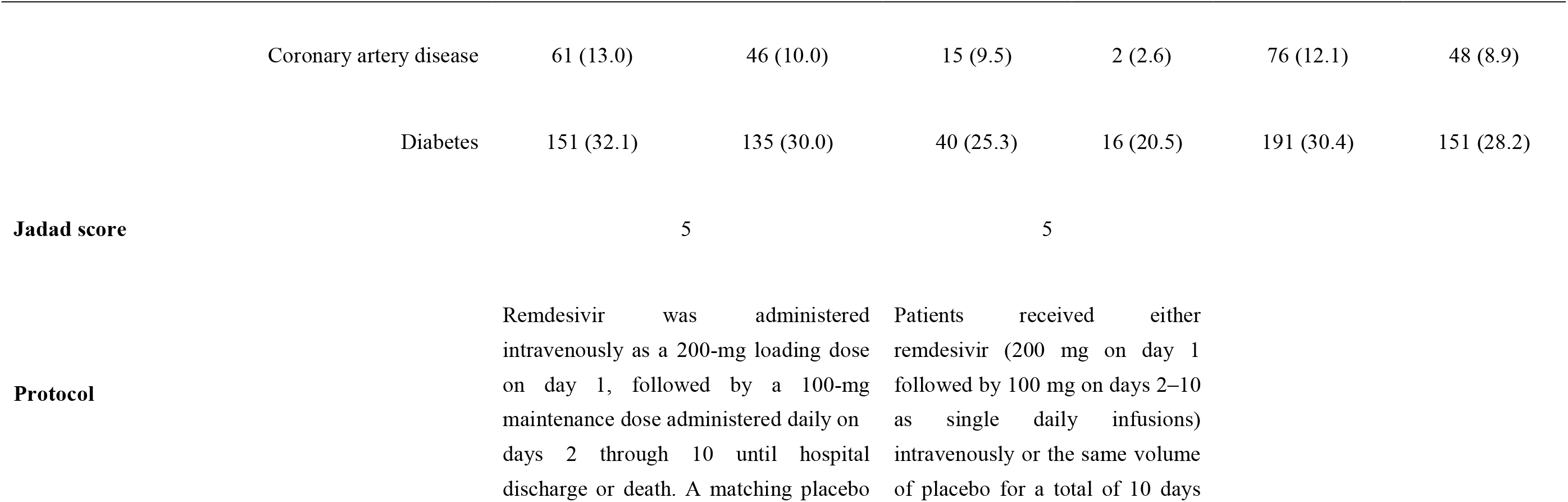
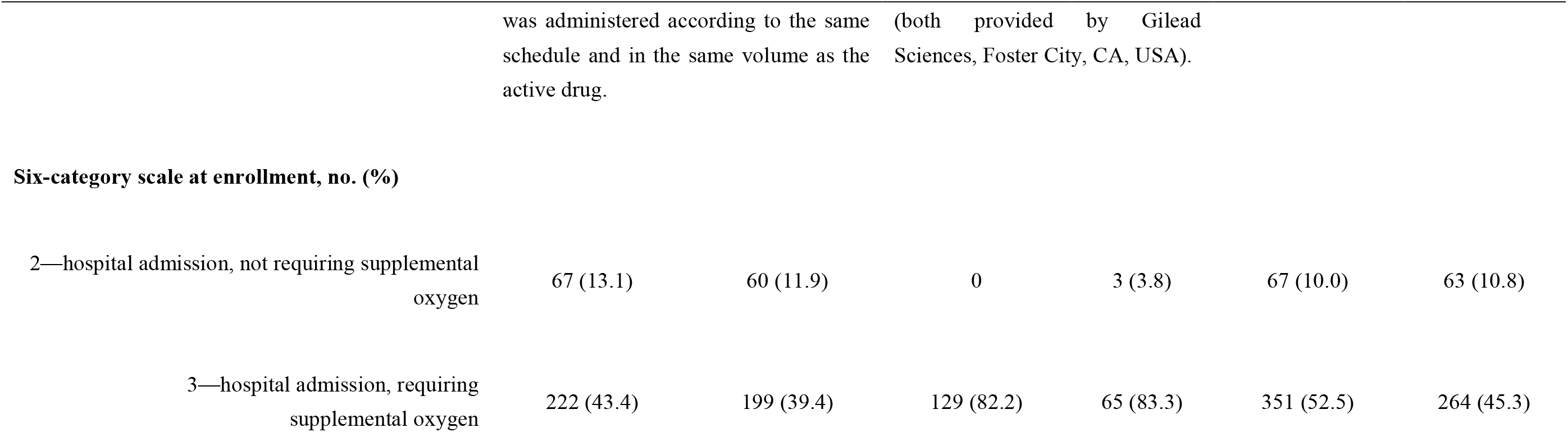
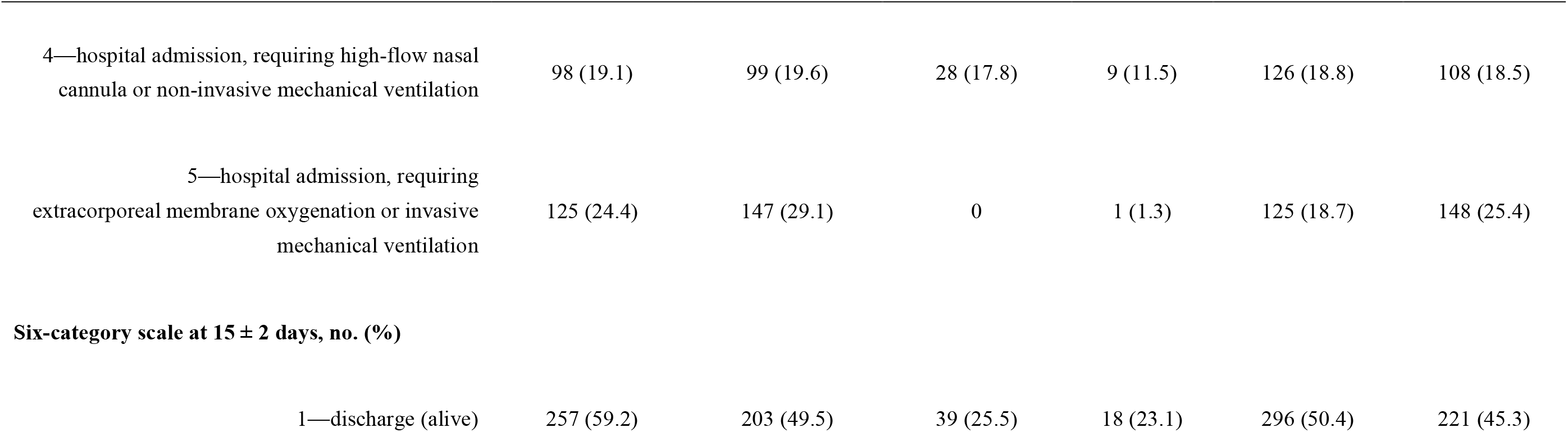
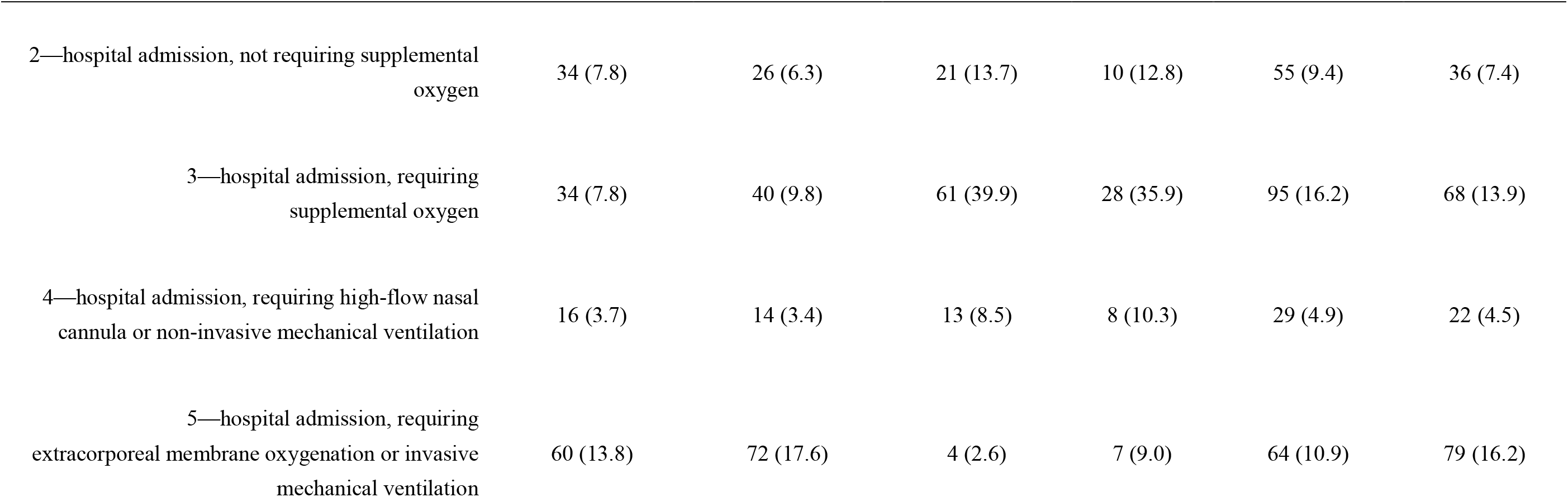
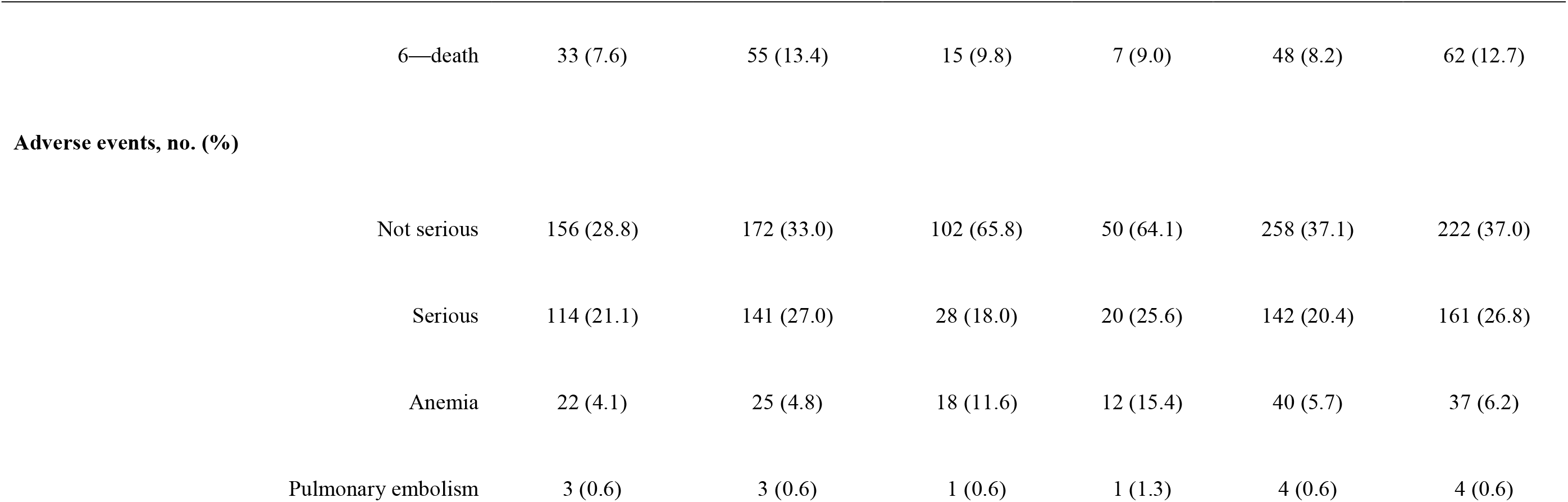
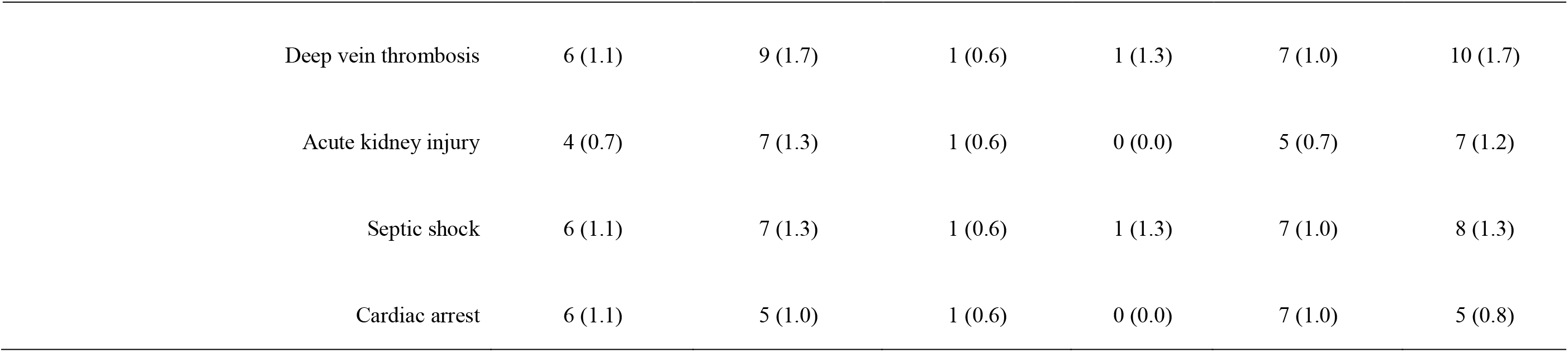
Study characteristics.

**Figure 1.**
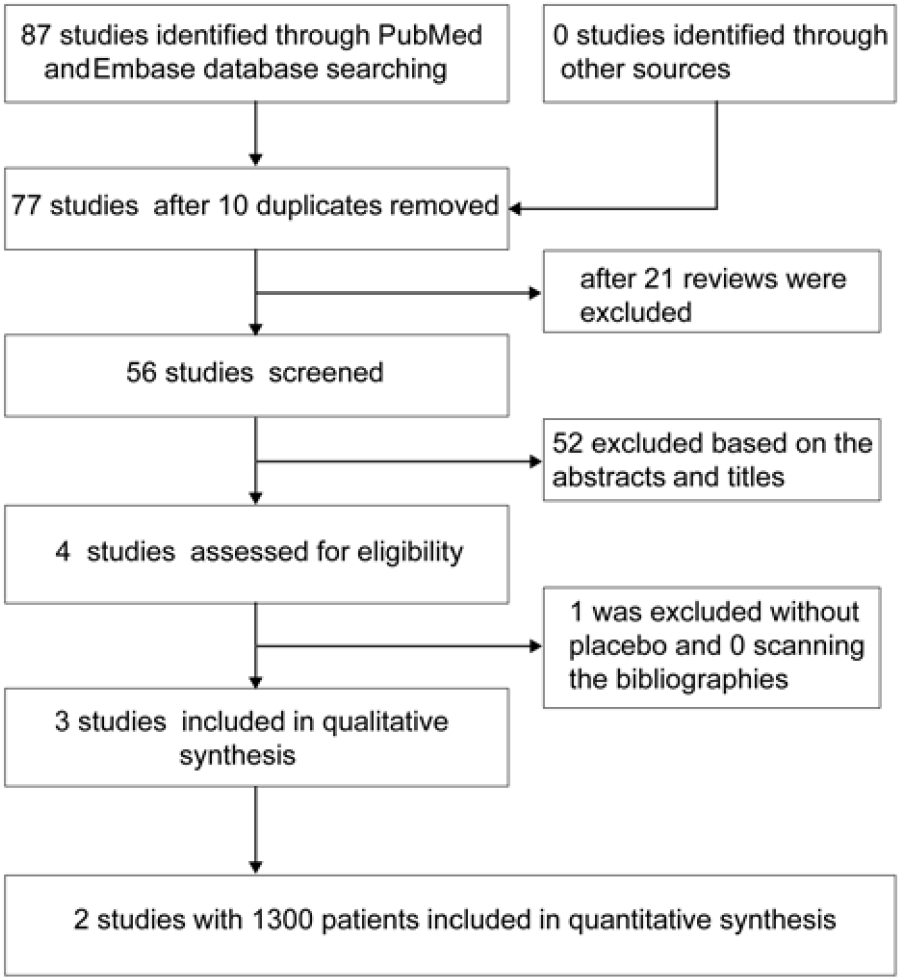
Flow diagram of study selection for pooled analysis.

**Figure 2.**
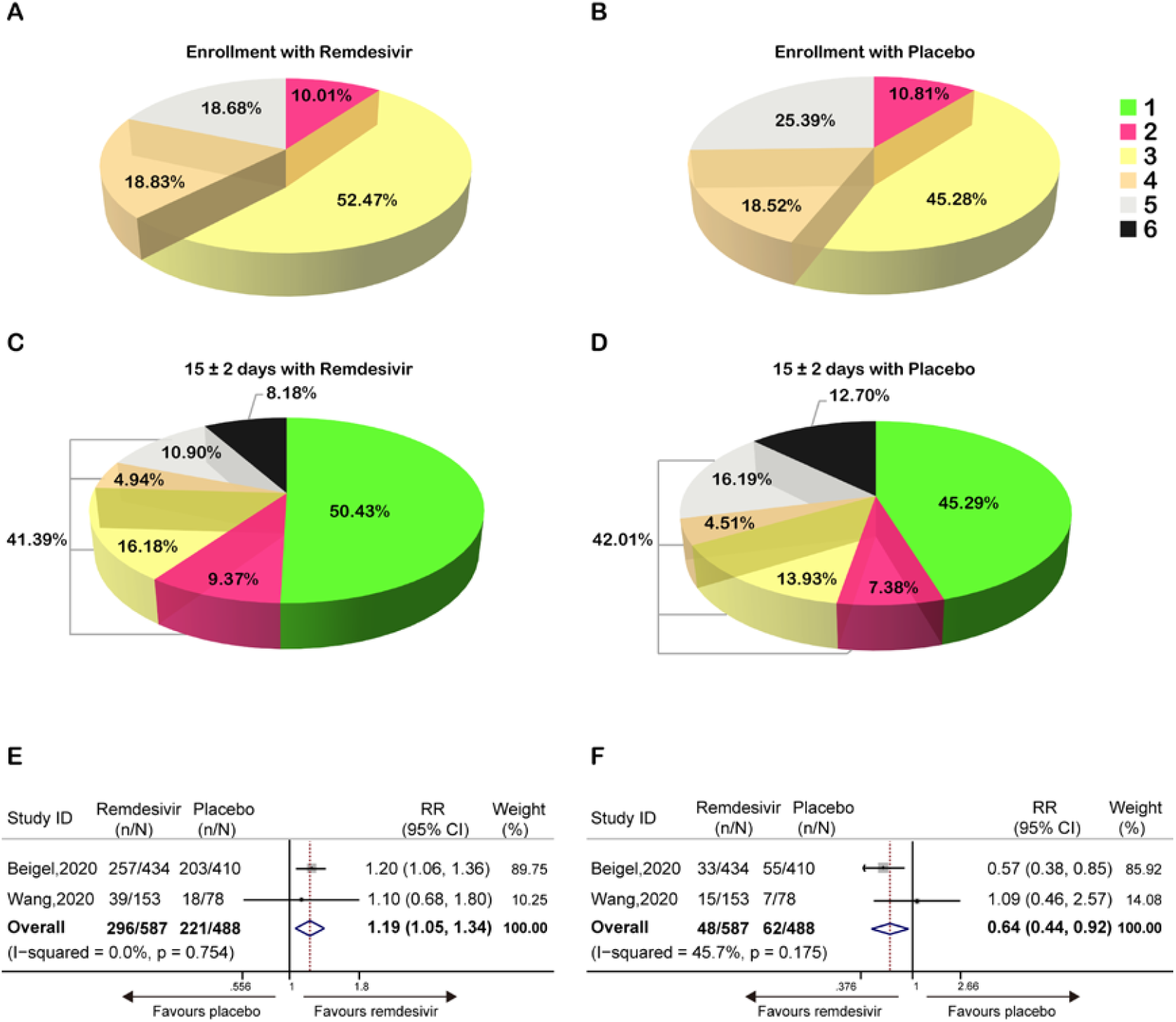
Treatment efficacy of remdesivir in patients with COVID-19 compared to the placebo based on a six-category scale at enrolment (1: Discharge (alive); 2: Hospital admission, not requiring supplemental oxygen; 3: Hospital admission, requiring supplemental oxygen; 4: Hospital admission, requiring high-flow nasal cannula or non-invasive mechanical ventilation; 5: Hospital admission, requiring extracorporeal membrane oxygenation or invasive mechanical ventilation; 6: Death). (A) Distribution of hospitalized patients with COVID-19 receiving remdesivir at enrollment. (B) Distribution of hospitalized patients with COVID-19 receiving placebo at enrollment. (C) Distribution of patients with COVID-19 receiving remdesivir at 15 ± 2 days. (D) Distribution of patients with COVID-19 receiving placebo at 15 ± 2 days. (E) Forest plots of estimated effects increasing hospital discharge of patients with COVID-19 receiving remdesivir compared with those receiving placebo. (F) Forest plots of the estimated effects for mortality in patients with COVID-19 receiving remdesivir compared with those receiving placebo.

First, among 1075 patients with COVID-19, patients in the remdesivir group had a higher rate of discharge and lower mortality than patients in the placebo group (50.43% vs. 45.29% and 8.18% vs. 12.70%, respectively) (Figure 2C, 2D). The pooled analysis indicated that remdesivir increased the discharge rate (RR, 1.19 [95% CI, 1.05–1.34]; P = 0·754, I^2^ = 0·0%) (Figure 2E) and decreased mortality (RR, 0.64 [95% CI, 0.44–0.92]; P = 0.175, I^2^ = 45.7%) (Figure 2F) in patients with COVID-19 compared with the placebo.

Second, the data of 1296 patients diagnosed with COVID-19 were analyzed to explore the association between remdesivir treatment and adverse events. The analysis suggested that remdesivir decreased the occurrence of serious adverse events (RR, 0.77 [95% CI, 0.63–0.94]; p = 0.716, I^2^ = 0.0%) (Figure 3A). Furthermore, remdesivir showed a tendency to reduce non-serious adverse events, although the tendency was not statistically significant (RR, 0.92 [95% CI, 0.80–1.06]; p = 0.225, I^2^ = 32.0%) (Figure 3B).

**Figure 3.**
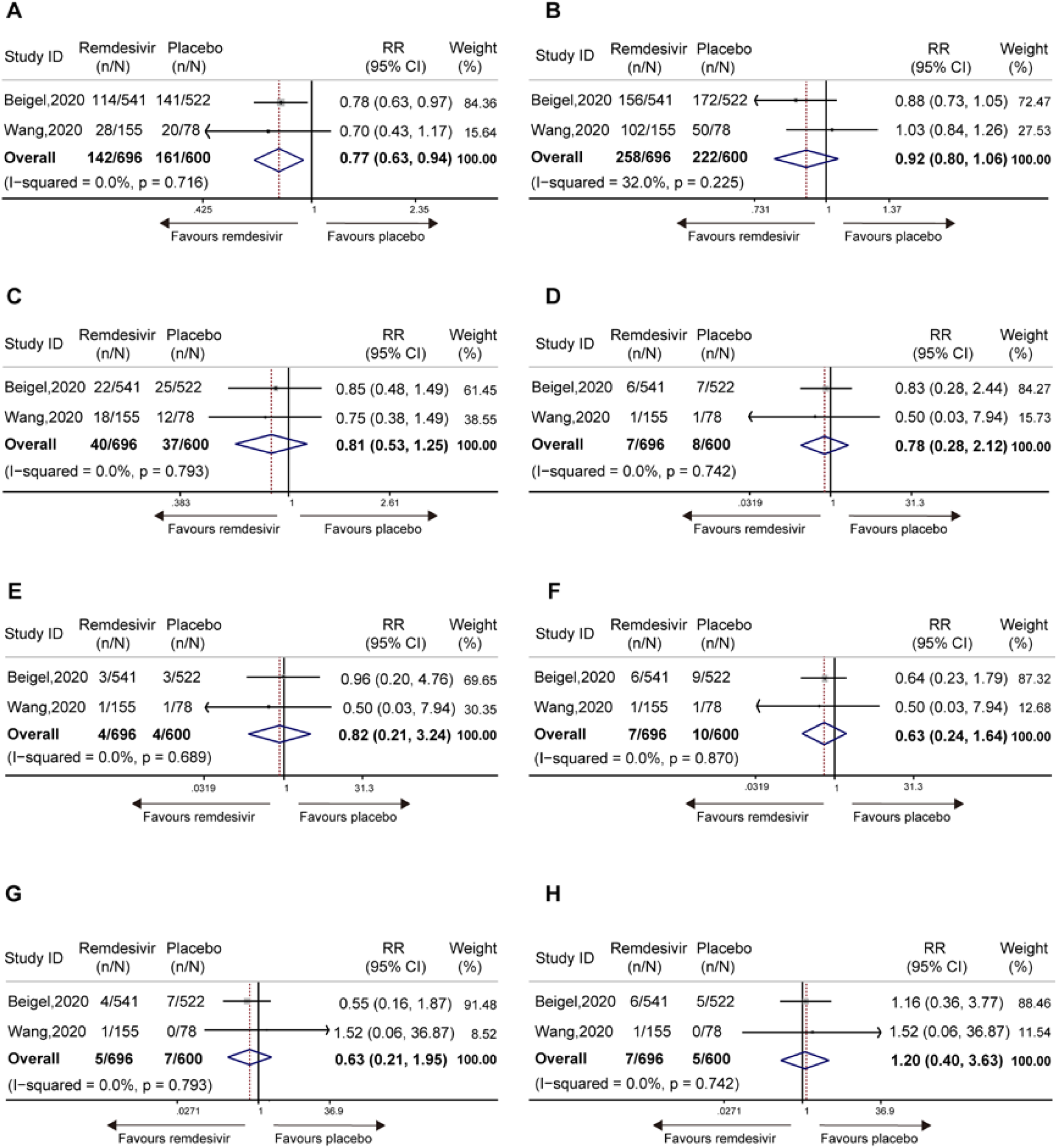
Forest plots of risk of adverse events for patients receiving remdesivir compared with the placebo. (A) Serious adverse events. (B) Non-serious adverse events. (C) Anemia. (D) Serious septic shock. (E) Serious pulmonary embolism. (F) Serious deep vein thrombosis. (G) Acute kidney injury. (H) Serious cardiac arrest.

Additionally, when anemia, septic shock, pulmonary embolism, deep vein thrombosis, acute kidney injury, and cardiac arrest were regarded as endpoints, remdesivir was associated with decreased risk of all of them, except cardiac arrest, although these differences were not statistically significant (Figure 3C-3H).

## Discussion

There is an urgent need for effective therapies for COVID-19. Remdesivir is a nucleotide analog prodrug with broad-spectrum antiviral activity and is currently being investigated in multiple COVID-19 clinical trials.^15^ Our analysis showed that remdesivir was effective and safe in treating patients with COVID-19.

First, our analysis indicated that remdesivir significantly increased the discharge rate of patients with COVID-19 at approximately 15 days (Figure 2A, 2B), indicative of its efficacy. Remdesivir has been reported to have potential benefits in severely ill patients with COVID-19.^10,16-17^ However, an open-label, randomized, multicenter trial in patients with severe COVID-19 presented a different result, that is, a non-significant difference between 5- and 10-day courses of remdesivir.^9^ Based on RCTs, Beigel et al. reported that patients treated with remdesivir had a shorter recovery time than those treated with placebo (median 11 days, compared to 15 days; RR for recovery, 1.32; 95% CI, 1.12–1.55; p < 0.001).^8^ A more rapid clinical improvement within 10 days was observed in patients receiving remdesivir compared to those receiving placebo (median 18·0 days [IQR 12.0–28.0] vs. 23.0 days [15.0–28.0]; HR 1.52 [0.95–2.43], P > 0.05).^7^ In our study, the discharge rate was used to more intuitively reflect the patient’s recovery or clinical improvement. The comprehensive analysis suggested that regardless of the proportion of patients in the initial six-category scale, the discharge rate in the remdesivir group was also higher than that in the placebo group, 50.43% vs. 45.29% (Table 1, Figure 2A, B). We believe that this result is more convincing than that presented by any single study.

Second, our results indicated that remdesivir significantly decreased the mortality of patients with COVID-19 (Figure 2C, 2D, 2F). Remdesivir has been shown to be effective in treating SARS-CoV-2 and related coronaviruses in an animal model;^18^ therefore, many studies have been conducted to test whether remdesivir can reduce mortality in patients with COVID-19. Two recent studies reported that remdesivir reduces mortality compared with placebo in patients with COVID-19 (8.0% vs. 11.6% and 7.1% vs. 11.9%),^8,19^ and this was consistent with our results (8.2% vs. 12.7%, Figure 2C, 2D). Furthermore, the mortality risk was 0.70 (95% CI, 0.47–1.04) in Beigel’s study,^8^ which presented results similar to ours (RR 0.64 [95% CI 0.44–0.92]); however, our result was statistically significant (Figure 2F). A study in China reported a different conclusion, with no difference in mortality between patients treated with remdesivir and placebo at 7, 14, and 28 days (6.5% vs. 5.2%, 9.8% vs. 9.0%, and 14.7% vs. 13.0%, respectively).^7^ However, its incomplete enrolment, owing to a lack of eligible patients and low persuasiveness, owing to its 2:1 randomization, may have made the experiment statistically inadequate. Because our study combined two high-quality randomized case-control studies, we believe that our results are more convincing. And previous vivo and vitro experiment also both supported our results. Remdesivir was found to prevent COVID-19 infection in vitro experiments at extremely low concentrations. ^5^ At the same time, a recent study found that remdesivir had clinical effect in rhesus monkeys infected with COVID-19: Rhesus monkeys treated early with reddisivir had less lung damage, no signs of respiratory infection, and less lower respiratory viral load than controls. ^15^

Third, our analysis indicated that remdesivir significantly decreased the risk of serious adverse events (Figure 3A) with a tendency to reduce the risk of non-serious adverse events (Figure 3B). Further evaluation of adverse events such as anemia, septic shock, pulmonary embolism, deep vein thrombosis, and acute kidney injury, showed similar trends. These data suggest that remdesivir is safe to be used for the treatment of COVID-19.

Our analysis had some limitations. First, the small number of studies that were included greatly weakened the reliability of our results. Second, the RR values calculated were unadjusted owing to a lack of raw data, which may lead to bias. Third, the overall sample size is not large enough, which limits statistical power. Thus, larger-scale RCTs are needed, and our pooled analysis findings should be interpreted with caution. However, we will update the analysis as new studies that meet inclusion criteria emerge.

Overall, this analysis demonstrated that remdesivir significantly increased the recovery rate, decreased mortality, and decreased the risk of serious adverse events. Our findings highlight the efficacy and safety of remdesivir for the treatment of COVID-19, which should be incorporated into routine therapy. Furthermore, this analysis also suggests that larger cohorts and randomized controlled trials are needed to more thoroughly investigate the treatment efficacy and safety of remdesivir for COVID-19.

## Data Availability

The data used to support the findings of this study are included within the article.

## Declaration of interests

We declare no competing interests.

## Acknowledgments

We thank all the contributors of the two eligible studies. We thank the Yunnan Key Laboratory of Digital Orthopaedics. We thank the National Natural Science Foundation of China (grants 81760136, 81660156, 31960136, and 81960268); the Joint Special Fund (grants 2018FE001(−175) and 2018FE001(−174)); and the Yunnan Health Training Project of High-level Talents (grants H-2017064 and H-2017028).

### 1. eAppendix 1: Literature search strategies

#### 1.1 PUBMED (20200611)

#1. remdesivir OR gs 5734

#2. ((((((COVID-19) OR (Coronavirus disease 2019)) OR (Novel Coronavirus 2019)) OR (2019 Novel Coronavirus)) OR (Severe acute respiratory syndrome-corona virus-2)) OR (SARS-CoV-2)) OR (Novel coronavirus pneumonia)

#3. (random*)OR “Randomized Controlled Trial” [pt] OR “Randomized Controlled Trials as Topic” [Mesh]

#4. #1 AND #2 AND #3

#### 1.2 Embase (20200611)

#1. ‘remdesivir’/exp OR remdesivir

#2. ‘gs 5734’/exp OR ‘gs 5734’ OR ((‘gs’/exp OR gs) AND 5734)

#3. #1 OR #2

#4. ‘covid 19’/exp OR ‘covid 19’ OR (covid AND 19)

#5. ‘coronavirus disease 2019’/exp OR ‘coronavirus disease 2019’ OR ((‘coronavirus’/exp OR coronavirus) AND (‘disease’/exp OR disease) AND 2019)

#6. ‘novel coronavirus 2019’ OR (novel AND (‘coronavirus’/exp OR coronavirus) AND 2019)

#7. ‘2019 novel coronavirus’/exp OR ‘2019 novel coronavirus’ OR (2019 AND novel AND (‘coronavirus’/exp OR coronavirus))

#8. ‘severe acute respiratory syndrome-corona virus-2’ OR (severe AND acute AND respiratory AND ‘syndrome corona’ AND ‘virus 2’)

#9. ‘sars cov 2’

#10. ‘novel coronavirus pneumonia’ OR (novel AND (‘coronavirus’/exp OR coronavirus) AND (‘pneumonia’/exp OR pneumonia))

#11. #4 OR #5 OR #6 OR #7 OR #8 OR #9 OR #10

#12. #3 AND #11

#13. crossover AND (‘procedure’/exp OR procedure) OR (double AND (‘blind’/exp OR blind) AND (‘procedure’/exp OR procedure)) OR (randomized AND controlled AND (‘trial’/exp OR trial)) OR (‘single blind’ AND (‘procedure’/exp OR procedure)) OR random* OR factorial* OR crossover* OR (cross AND over*) OR placebo* OR (doubl* AND adj AND blind*) OR (singl* AND adj AND blind*) OR assign* OR allocat* OR volunteer*

#14. #12 AND #13

#### 1.3 Cochrane Central Register of Controlled Trials (20200611)

(Remdesivir OR GS-5734) AND (COVID-19 OR Coronavirus disease 2019 OR Novel Coronavirus 2019 OR Novel Coronavirus 2019 OR 2019 Novel Coronavirus OR Severe acute respiratory syndrome-corona virus-2 OR SARS-CoV-2 OR Novel coronavirus pneumonia)

### 2. eAppendix 2: The six-point scale categories^1^

| Ordinal Scale | Characteristic |
| --- | --- |
| 1. | not hospitalized |
| 2. | hospitalized, not requiring supplemental oxygen |
| 3. | hospitalized, requiring supplemental oxygen |
| 4. | hospitalized, requiring nasal high-flow oxygen therapy, noninvasive mechanical ventilation, or both |
| 5. | hospitalized, requiring invasive mechanical ventilation, ECMO, or both |
| 6. | death |

1. Wang Y, Zhang D, Du G, et al. Remdesivir in adults with severe COVID-19: a randomised, double-blind, placebo-controlled, multicentre trial. Lancet 2020;395:1569–78.

## Notes

### Competing Interest Statement

The authors have declared no competing interest.

### Clinical Trial

This study is registered at INPLASY (INPLASY202060046).

### Funding Statement

This study was supported by the Joint Special Fund (grants 2018FE001(-175) and 2018FE001(-174)); and the Yunnan Health Training Project of High-level Talents (grants H-2017064 and H-2017028).

